## Supplementary Information for "Tracing contacts to evaluate the transmission of COVID-19 from highly exposed individuals in public transportation"

(Dated: May 31, 2021)

### I. EPIDEMIOLOGICAL CURVES FOR THE COMPARTMENTAL MODEL

We show all epidemiological curves for the proposed compartmental model in Fig. S1a-S1f. The curve of the susceptible population is shown as a percentage. We estimate that about 20% of the inhabitants of Fortaleza had contact with SARS-CoV-2 until December 2020. For the deceased population, we show the comparison between the observed cumulative number of deaths by SARS-CoV-2 in Fortaleza and our inferred model prediction.

The time series of the effective reproduction number $Re^{city}$ for several values of $\alpha$ is shown in Fig. S2. We find that $Re^{city}$ remains practically the same for $\alpha = 0.15$ and 0.20, which are compatible with the values of fractions of reported infectious individuals previously reported in the literature [1]. On March 20, 2020, the Government of Ceará imposed the first social distancing decree to its population, which led to a substantial decrease in human mobility everywhere in the state. However, as depicted, a value of $Re^{city}$ around 2 in the beginning of April still indicated a rapid and exponential-like increase of the epidemic in the city of Fortaleza. A steep decrease in $Re^{city}$ was then observed and that effective reproduction number dropped below the critical value 1 around May 5, 2020, when the Government decreed lockdown. The regime of lockdown imposed to the population of Fortaleza, in which only the commuting of essential workers were allowed, was responsible to keep the virus transmission under control ($Re^{city} < 1$) until at least October 2020.

### II. TIME EVOLUTION OF THE UPSCALING FACTOR

The time evolution of the upscaling factor $\chi/\psi$ is shown in Fig. S3.

### III. ITERATED ENSEMBLE KALMAN FILTER FRAMEWORK

As shown in Fig. S4, the flowchart of the Iterated Ensenble Kalman Filter (IEnKF) [1–4] employed in our work includes the solution of the SEIIR model as a core modulus fed with initial guesses for state and parameter vectors. This modulus is built within the inference algorithm, from which estimated parameters and states during a given time window are obtained.

### IV. PARAMETERS OF THE COMPARTMENTAL MODEL

Table S1 shows the epidemiological parameters of SEIIR model adopted for each window of inference.

---


[1] Li, R. *et al.* Substantial undocumented infection facilitates the rapid dissemination of novel coronavirus (SARS-CoV-2). *Science* **368**, 489-493 (2020). https://doi.org/10.1126/science.abb3221

[2] Ionides, E. L., Bretó, C. & King, A. A. Inference for nonlinear dynamical systems. *Proc. Natl. Acad. Sci. U. S. A.* **103**, 18438-18443 (2006). https://doi.org/10.1073/pnas.0603181103

[3] King, A. A., Ionides, E. L., Pascual, M. & Bouma, M. J. Inapparent infections and cholera dynamics. *Nature* **454**, 877-880 (2008). https://doi.org/10.1038/nature07084

[4] Sakov, P., Oliver, D. S. & Bertino, L. An Iterative EnKF for Strongly Nonlinear Systems. *Monthly Weather Review* **140**, 1988–2004 (2012). https://doi.org/10.1175/MWR-D-11-00176.1

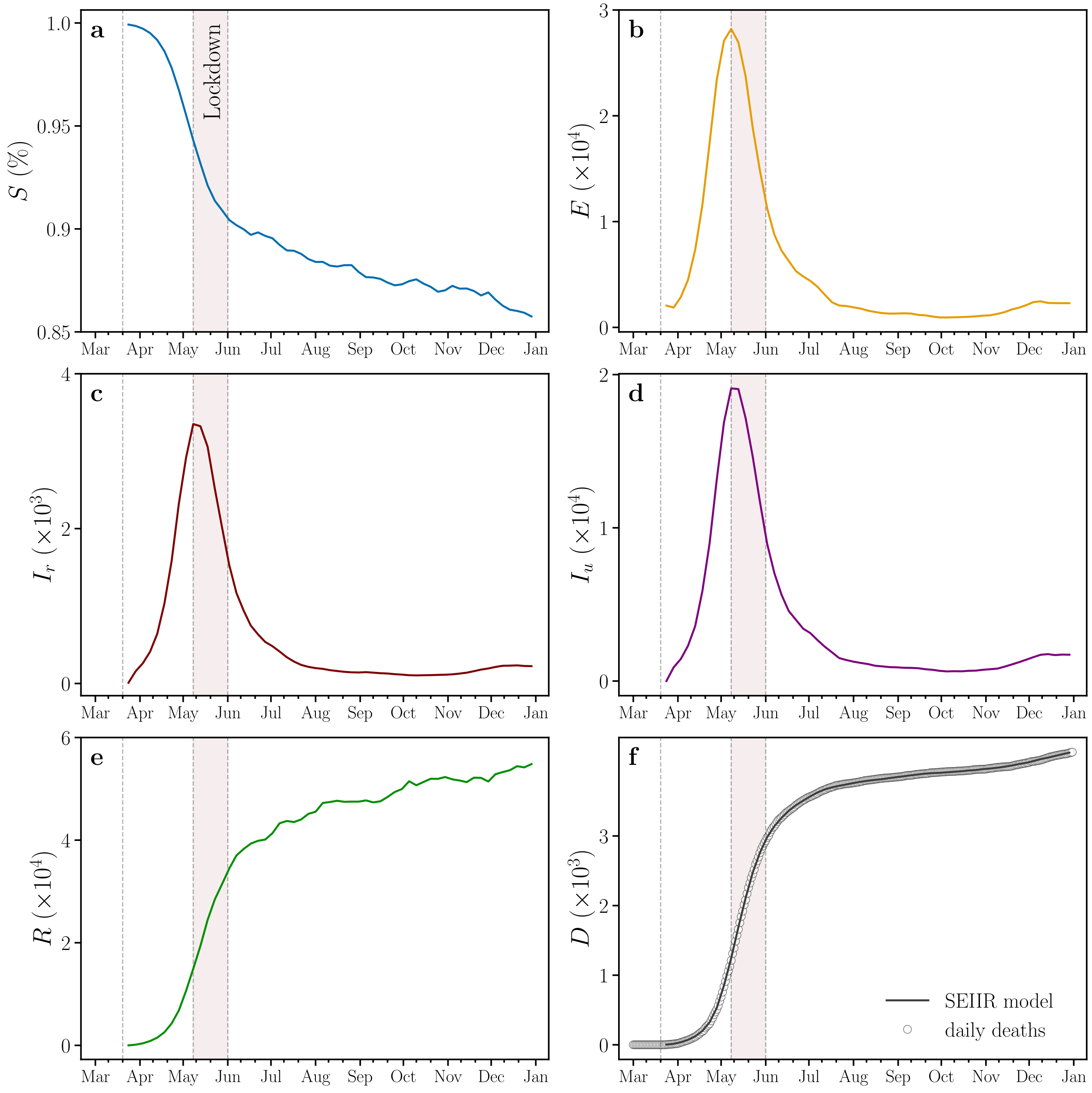


Fig. S1. **Epidemiological curves obtained from the inference with the SEIIR compartmental model.** Curves of the SEIIR model compartments. We show the curves with the values of the (a) susceptible, (b) exposed, (c) reported infectious, (d) unreported infectious, (e) recovered and (f) deceased population, respectively. The vertical dotted lines represent the beginning of social isolation (State Decree 33,519), lockdown (State Decree 33,574), and economic reopening (State Decree 33,608) regimes imposed on March 20, May 8, and June 1, 2020, respectively. We also highlight, in light red, the lockdown period in the city of Fortaleza.

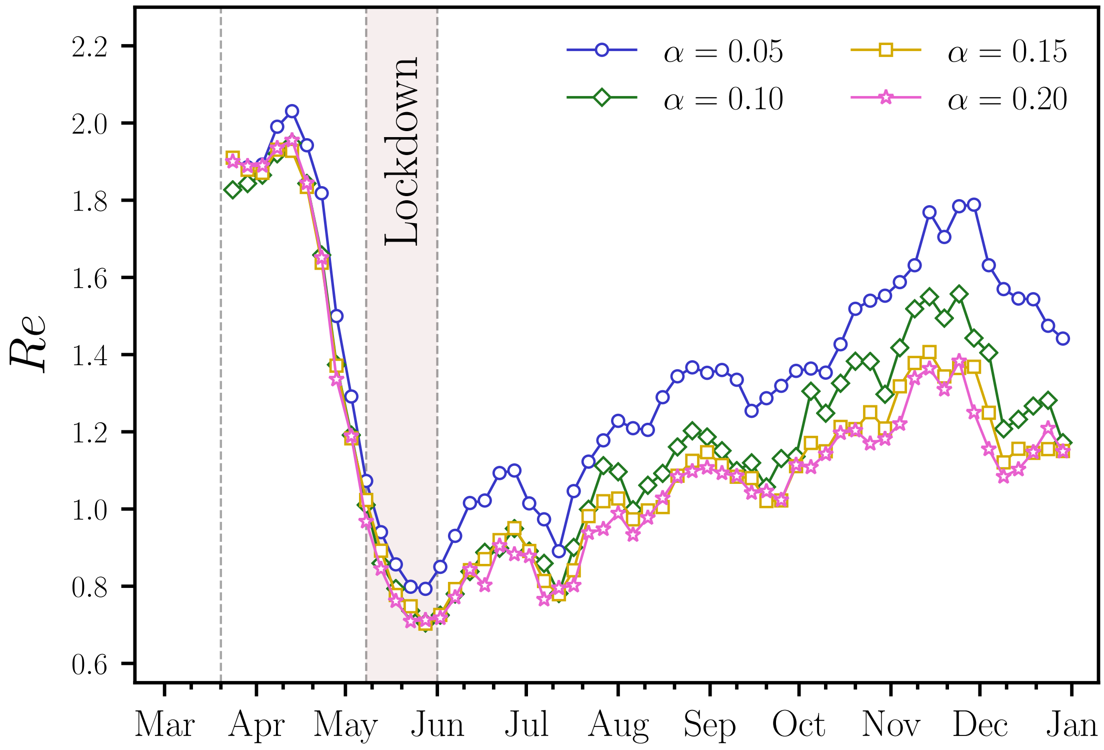


Fig. S2. **Time Evolution of the effective reproduction number for the entire city**. We show the $Re^{city}$ curve computed from the SEIIR model for $\alpha = 0.055$, 0.10, 0.15 and 0.20. Each point represents a bin of 22 days width and a 5 days step size. The vertical dotted lines represent the beginning of social isolation (State Decree 33,519), lockdown (State Decree 33,574), and economic reopening (State Decree 33,608) regimes imposed on March 20, May 8, and June 1, 2020, respectively. We also highlight, in light red, the lockdown period in the city of Fortaleza.

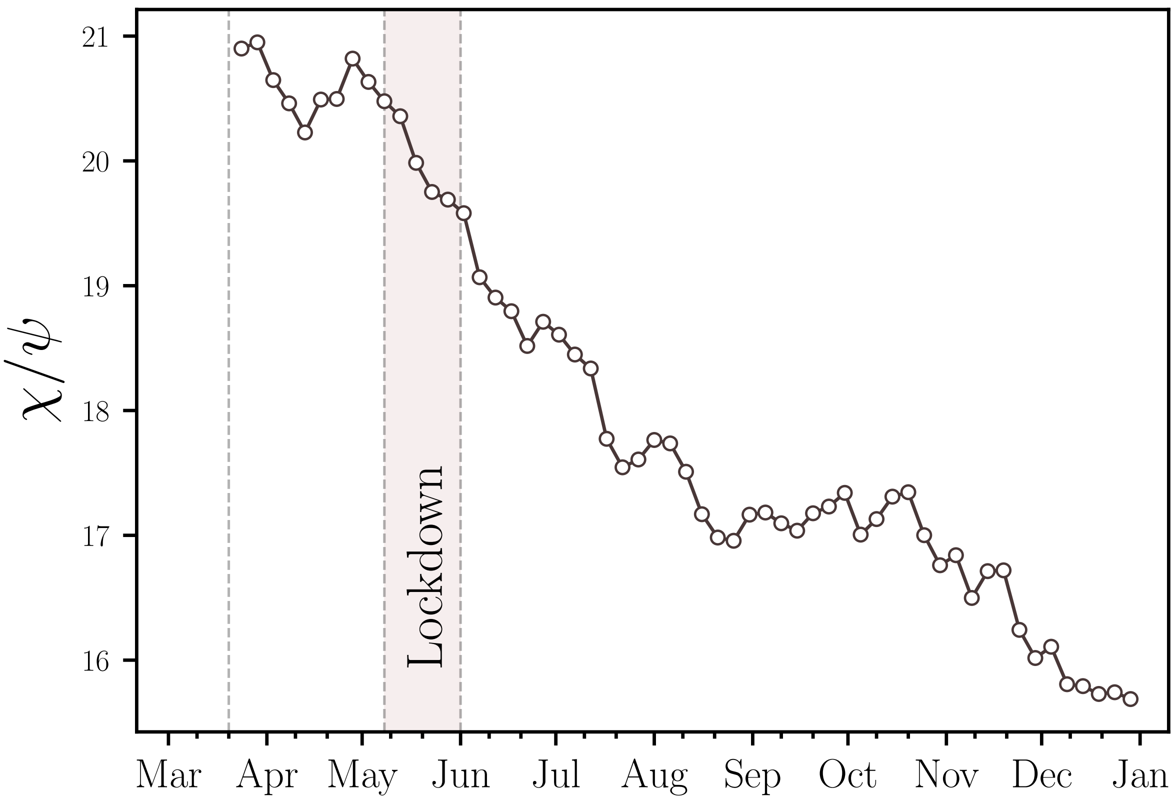


Fig. S3. **Time Evolution of the upscaling factor**. The value of the upscaling factor $\chi/\psi$ computed from the SEIIR model decreases for consecutive time windows. Each point represents a bin of 22 days width and a 5 days step size. The vertical dotted lines represent the beginning of social isolation (State Decree 33,519), lockdown (State Decree 33,574), and economic reopening (State Decree 33,608) regimes imposed on March 20, May 8, and June 1, 2020, respectively. We also highlight, in light red, the lockdown period in the city of Fortaleza.

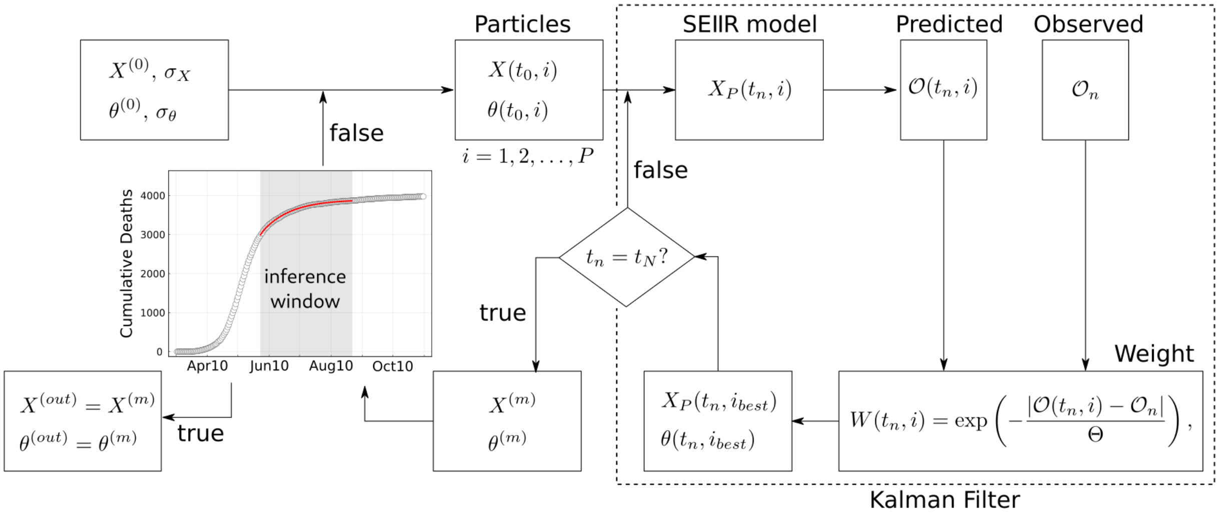


Fig. S4. **Iterated Ensemble Kalman Filter (IEnKF) Framework.** Step-by-step flowchart of the IEnKF used for the inference with the compartmental model.

| Date | $\beta$ | $\mu$ | $\alpha$ | $\sigma$ | $\gamma$ | $\phi$ |
|---|---|---|---|---|---|---|
| 2020-03-24 | 0.941742 | 0.486745 | 0.149977 | 0.223059 | 0.281187 | 0.069320 |
| 2020-03-29 | 0.923219 | 0.486771 | 0.152547 | 0.221021 | 0.281529 | 0.069493 |
| 2020-04-03 | 0.937314 | 0.476879 | 0.152980 | 0.221588 | 0.283996 | 0.067959 |
| 2020-04-08 | 0.971974 | 0.471830 | 0.151557 | 0.218856 | 0.284957 | 0.066534 |
| 2020-04-13 | 0.975671 | 0.464336 | 0.151683 | 0.222570 | 0.281936 | 0.066516 |
| 2020-04-18 | 0.913568 | 0.471593 | 0.153497 | 0.220646 | 0.285345 | 0.067107 |
| 2020-04-23 | 0.818039 | 0.472312 | 0.152583 | 0.225127 | 0.285158 | 0.071127 |
| 2020-04-28 | 0.685570 | 0.480382 | 0.156198 | 0.231284 | 0.296786 | 0.075009 |
| 2020-05-03 | 0.601887 | 0.472099 | 0.159905 | 0.232949 | 0.306464 | 0.076051 |
| 2020-05-08 | 0.531756 | 0.467665 | 0.159058 | 0.232452 | 0.314061 | 0.078509 |
| 2020-05-13 | 0.476636 | 0.464158 | 0.158516 | 0.234426 | 0.318556 | 0.077596 |
| 2020-05-18 | 0.429962 | 0.452728 | 0.157680 | 0.235465 | 0.321578 | 0.078138 |
| 2020-05-23 | 0.418717 | 0.445415 | 0.157436 | 0.232583 | 0.315319 | 0.076398 |
| 2020-05-28 | 0.393294 | 0.444844 | 0.155333 | 0.230803 | 0.316501 | 0.076914 |
| 2020-06-02 | 0.410878 | 0.443603 | 0.151952 | 0.228969 | 0.315920 | 0.076274 |
| 2020-06-07 | 0.460817 | 0.428248 | 0.150497 | 0.232291 | 0.315902 | 0.075106 |
| 2020-06-12 | 0.496968 | 0.424960 | 0.147721 | 0.227823 | 0.313617 | 0.074771 |
| 2020-06-17 | 0.520876 | 0.423615 | 0.144601 | 0.228621 | 0.314275 | 0.073622 |
| 2020-06-22 | 0.553313 | 0.415472 | 0.143676 | 0.222961 | 0.313110 | 0.073552 |
| 2020-06-27 | 0.563638 | 0.422421 | 0.142414 | 0.220978 | 0.315451 | 0.071979 |
| 2020-07-02 | 0.537569 | 0.420991 | 0.139717 | 0.220920 | 0.316575 | 0.071163 |
| 2020-07-07 | 0.492326 | 0.414203 | 0.142366 | 0.221908 | 0.313338 | 0.069663 |
| 2020-07-12 | 0.472757 | 0.410928 | 0.142010 | 0.226979 | 0.313961 | 0.067919 |
| 2020-07-17 | 0.529636 | 0.394527 | 0.140153 | 0.224913 | 0.312099 | 0.067265 |
| 2020-07-22 | 0.623454 | 0.389191 | 0.137581 | 0.221728 | 0.310518 | 0.065810 |
| 2020-07-27 | 0.651591 | 0.391906 | 0.136511 | 0.224862 | 0.309801 | 0.065467 |
| 2020-08-01 | 0.646985 | 0.394598 | 0.139634 | 0.229596 | 0.310438 | 0.066236 |
| 2020-08-06 | 0.619746 | 0.394033 | 0.139218 | 0.229674 | 0.312464 | 0.065691 |
| 2020-08-11 | 0.636491 | 0.386854 | 0.139302 | 0.230407 | 0.309448 | 0.065631 |

| | | | | | | |
|---|---|---|---|---|---|---|
| 2020-08-16 | 0.649350 | 0.374605 | 0.141486 | 0.231604 | 0.308376 | 0.066573 |
| 2020-08-21 | 0.700571 | 0.369588 | 0.140327 | 0.231880 | 0.302333 | 0.067226 |
| 2020-08-26 | 0.728535 | 0.368281 | 0.140994 | 0.230835 | 0.301127 | 0.066636 |
| 2020-08-31 | 0.737557 | 0.374409 | 0.141605 | 0.228592 | 0.296083 | 0.067373 |
| 2020-09-05 | 0.706375 | 0.375461 | 0.140895 | 0.226542 | 0.295648 | 0.067289 |
| 2020-09-10 | 0.691229 | 0.374577 | 0.138387 | 0.226096 | 0.295912 | 0.067328 |
| 2020-09-15 | 0.690880 | 0.372604 | 0.138551 | 0.227870 | 0.291531 | 0.067811 |
| 2020-09-20 | 0.648453 | 0.375282 | 0.140883 | 0.232778 | 0.289339 | 0.066958 |
| 2020-09-25 | 0.650962 | 0.375601 | 0.142775 | 0.231963 | 0.292576 | 0.067333 |
| 2020-09-30 | 0.705538 | 0.378565 | 0.143436 | 0.231854 | 0.298863 | 0.066841 |
| 2020-10-05 | 0.758124 | 0.370228 | 0.140453 | 0.233296 | 0.298339 | 0.067408 |
| 2020-10-10 | 0.742961 | 0.374269 | 0.140271 | 0.229163 | 0.296711 | 0.066331 |
| 2020-10-15 | 0.766765 | 0.380999 | 0.138758 | 0.229484 | 0.292138 | 0.066040 |
| 2020-10-20 | 0.763361 | 0.381141 | 0.140105 | 0.228106 | 0.297784 | 0.067130 |
| 2020-10-25 | 0.802157 | 0.370811 | 0.139463 | 0.228721 | 0.302356 | 0.067226 |
| 2020-10-30 | 0.782503 | 0.361956 | 0.141222 | 0.225585 | 0.304570 | 0.066343 |
| 2020-11-04 | 0.845170 | 0.365497 | 0.139852 | 0.229970 | 0.301449 | 0.065995 |
| 2020-11-09 | 0.911679 | 0.356743 | 0.137196 | 0.232050 | 0.308682 | 0.064187 |

Tab. S1: **Parameters of the compartmental model.** The epidemiological parameters of SEIIR model adopted for each window of inference.
